# Serum Glial Fibrillary Acidic Protein: Potential to Determine Stroke-Type, Time-Line and Tissue-Impact in Acute Stroke

**DOI:** 10.1101/2024.07.10.24310119

**Authors:** Julien F. Paul, Célina Ducroux, Pamela Correia, Audrey Daigneault, Catherine Larochelle, Christian Stapf, Laura C. Gioia

## Abstract

**Background:** Interest is emerging regarding the role of blood biomarkers in acute stroke. The aim of this pilot study was to determine the feasibility of biomarker acquisition in suspected acute stroke, using modern ultrasensitive immunoassay techniques, and explore their potential usefulness for stroke diagnosis and management.

**Methods:** In 62 patients with suspected acute stroke, blood samples were prospectively obtained upon arrival and prior to neuroimaging. Serum levels (pg/mL) of glial fibrillary acidic protein (GFAP) and neurofilament light chain (NfL) were analyzed according to time of symptom onset, neuroimaging, and final diagnosis.

**Results:** Acute ischemic stroke (AIS) was diagnosed in 35 patients, 10 with large-vessel occlusion (LVO). The remaining were diagnosed with intracerebral hemorrhage (ICH) (n = 12), transient ischemic attack (n=4), and stroke mimics (n=11). Median(IQR) GFAP levels were significantly higher in ICH (2,877.8 [1,002.1-10,402.5] pg/mL) compared to others diagnoses. In AIS, GFAP levels appear to increase over time from symptom onset, and were higher in patients with more extensive ischemic changes on baseline CT (ASPECTS ≤ 7) than those without, particularly in LVO stroke. NfL values were similar across groups.

**Conclusions:** In acute stroke, serum GFAP shows potential as an adjunct tool for the distinction between ICH and AIS. Specific to AIS, GFAP may also offer insight into time from onset, and extent of ischemic tissue injury on neuroimaging, particularly in LVO stroke. These preliminary findings merit further study.

## INTRODUCTION

Current management of acute ischemic stroke (AIS) and intracerebral hemorrhage (ICH), requires 1) neuroimaging to provide a diagnosis and 2) a time of symptom onset to determine eligibility for treatment. In up to 30% of patients, however, time of symptom onset is either unknown or exceeds recommended time windows for treatment.^(1)^ Advanced neuroimaging biomarkers are now widely recognized key criteria in acute stroke management, independent of time, to identity eligible patients.^(2)^ Accordingly, a paradigm shift is evolving away from time-based decision algorithms and toward physiology-based acute stroke management strategies. By extension, brain-specific blood biomarkers may offer an innovative opportunity to provide physiology-based information and potentially improve accessibility to acute stroke treatments.

Glial fibrillary acidic protein (GFAP), an astrocytic protein found almost exclusively in the brain, is a promising biomarker of brain tissue damage in neurological conditions, including stroke.^(3)^ In the first 6 hours, GFAP can discriminate between AIS and ICH,^(4)^ presumably due to acute brain damage incurred following hematoma formation and immediate disturbance of the brain blood barrier in ICH compared to AIS where there is a slower transition between penumbral tissue into core over time in the absence of reperfusion.^(5)^ Neurofilament light chain (NfL), a novel biomarker for axonal injury, shows promise as a prognostic tool following AIS.^(6)^ However, data regarding its use in acute stroke is limited. In previous studies, GFAP was measured using ELISA techniques (expressed in ng/mL) or did not include stroke patients beyond the hyperacute phase, thus limiting information in acute stroke of unknown or delayed onset. The aim of this pilot study was to evaluate the feasibility of blood biomarker acquisition upon Emergency Department (ED) arrival in suspected acute stroke using a novel ultrasensitive immunoassay (SIMOA). In addition, we conducted exploratory analyses to compare biomarker levels according to time of symptom onset, acute neuroimaging, and final diagnosis.

## METHODS

In this prospective pilot study, patients with suspected acute stroke (<24 hours from known symptom onset or last seen well) were recruited over 6 months. Blood samples were obtained during routine blood draw upon ED arrival. After 30 minutes to allow for coagulation, samples were centrifuged at 1800g for 10 minutes at 15-24ºC to separate cells and serum. Serum was then frozen at −80°C in aliquoted cryotubes. Serum GFAP and NfL levels were measured in duplicates with a SR-X ultra-sensitive biomarker detection system and commercially available Neurology 2-Plex B assay. Intra-assay variabilities (% coefficient of variation, CV) for duplicate measures was 6.2% (GFAP) and 6.9% (NfL); 88% of samples had intra-assay %CV <20%. Biomarker levels were compared according to final diagnosis (AIS, ICH, transient ischemic attack (TIA), or stroke mimic), time from onset, and neuroimaging (large-vessel occlusion (LVO) stroke presence, ASPECTS score). The protocol was approved by the institutional ethics committee. Patient or legal representative consent was obtained for study inclusion. Anonymized data was collected and stored in an encrypted database and extracted for analysis using R software. The corresponding author has full access to study data, and takes responsibility for study integrity and data analysis.

### Statistical Analyses

GFAP and NfL values among groups were compared using a Kruskall-Wallis test, followed by Dunn’s post-hoc test. Correlation between time of stroke onset and GFAP values was performed using Spearman rho correlation coefficient and stratified for the presence of LVO. Comparisons between poor (≤7) and favourable ASPECTS (8-10) were made using Wilcoxon Rank Sum test.

## RESULTS

Blood samples at ED arrival were collected in 68 patients, though 6 patients were excluded since blood sample processing times were delayed. Among the remaining 62 patients, median (IQR) time from ED presentation to blood draw was 11 (9-24) minutes and 48 (37-70) minutes from sample acquisition to centrifugation. AIS was diagnosed in 35 patients, of which 10 had a LVO, ICH (n=12), TIA (n=4), and stroke mimics (n=11). Baseline characteristics are summarized in Table 1. Acute stroke patients (AIS or ICH) had higher median GFAP concentrations (384.9 [164.25, 1,576.63] pg/mL) compared to those with TIA or stroke mimics (117.8 [65.8, 307.4] pg/mL) (p = 0.002), with ICH showing highest median GFAP levels (2,877.8 [1,002.1-10,402.5] pg/mL). (Figure 1A) NfL levels were similar between groups. (Figure 1B)

**Table 1:** Baseline patient characteristics.

|  | Overall (n=62) |
| --- | --- |
| Age, years (mean±SD) | 68.5±16.5 |
| Females, n(%) | 27 (43.5) |
| modified Rankin Score, median[IQR] | 1 [0 - 1] |
| Initial NIHSS, median[IQR] | 9 [3 - 17] |
| Time. stroke onset-ED arrival (minutes), median(IQR) |  |
| - Known onset | 82 [53-120] |
| - Unknown onset (last seen well) | 690 [277-870] |
| Patients with unknown onset, n(%) | 31 (50) |
| Time, ED arrival-sample acquisition (minutes) | 11 [9-24] |
| Time, sample acquisition-centrifugation (minutes) | 48 [37-70] |
| ***Final Diagnosis*** |  |
| ***Acute ischemic stroke*** | 35 |
| - Large vessel occlusion (LVO) | 10 |
| - ASPECTS score, median [IQR] | 10 [9-10] |
| - ASPECTS ≤ 7 | 5 |
| ***Intracerebral hemorrhage*** | 12 |
| ***Transient Ischemic Attack*** | 4 |
| ***Stroke Mimic*** | 11 |

**Figure 1:**
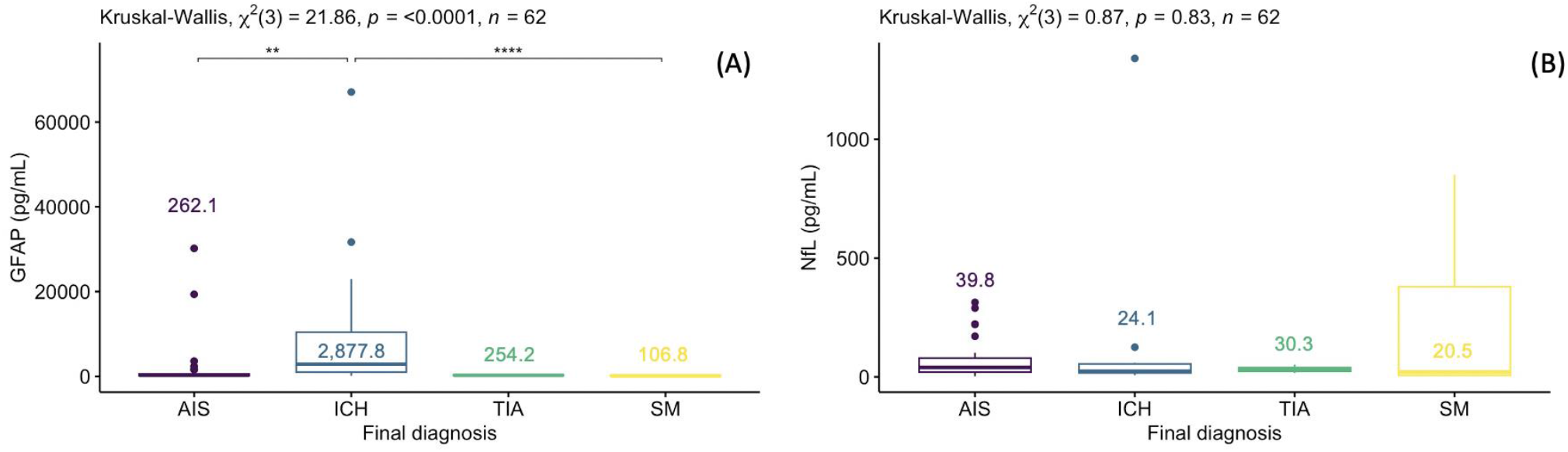
Box plot with median values of GFAP (A) and NfL (B) values in pg/mL in patients presenting with suspected stroke, classified according to their final diagnosis

In AIS with known time of stroke onset, GFAP levels appeared to increase linearly over time, particularly in LVO stroke (R=0.9, p=0.08). (Figure 2A) Furthermore, median GFAP levels appeared higher in AIS with more extensive ischemic changes on baseline CT (defined a priori as ASPECTS ≤ 7) (1578.87 [239.78, 2370.47] pg/mL) than those without (246.71 [135.52, 444.54] pg/mL) (Fig.2B), particularly when stratified by LVO presence. (Fig.2C) No association was found between NfL and neuroimaging or time from stroke onset.

**Figure 2:**
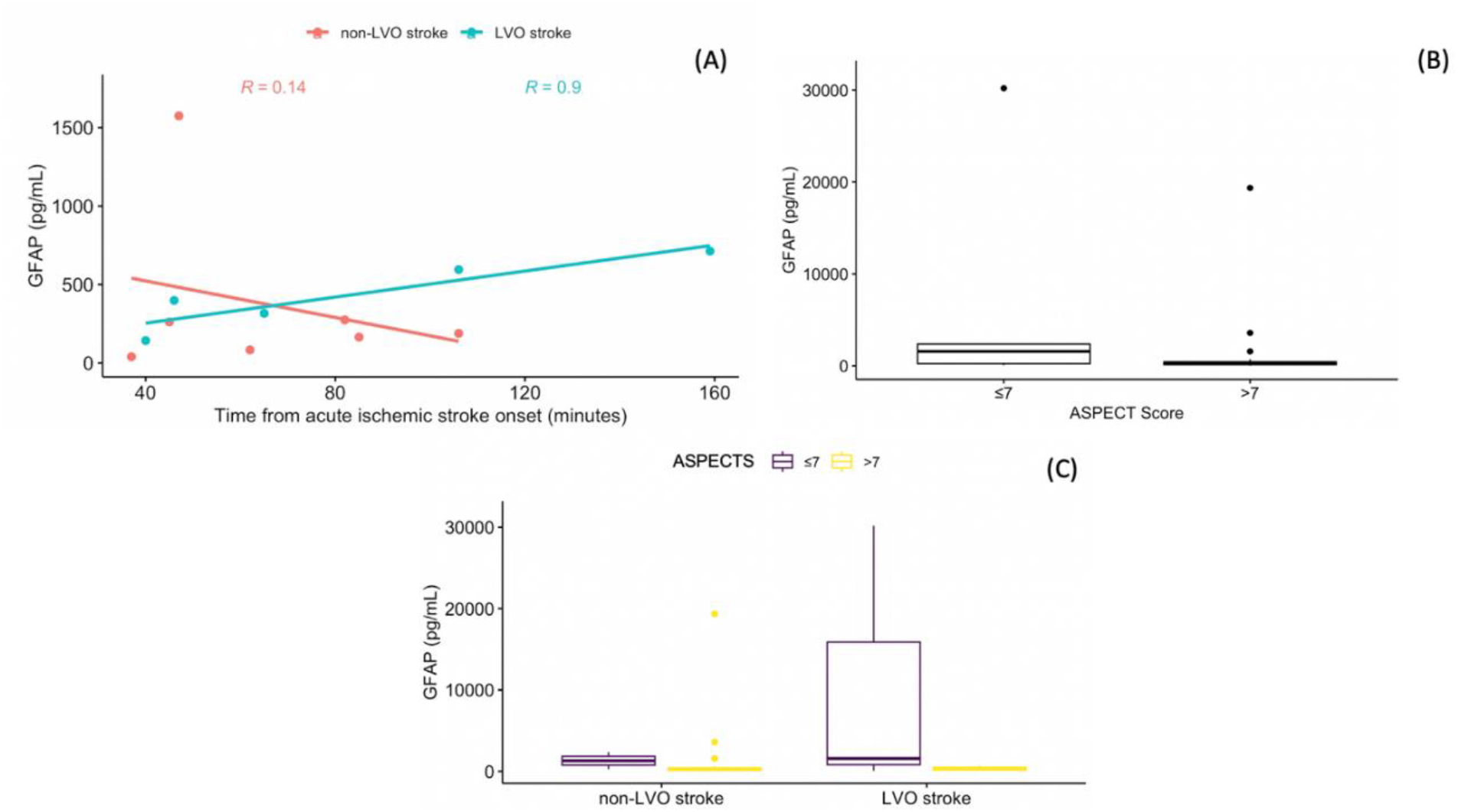
Scatterplots with regression lines of GFAP in AIS with known onset (A), stratified for LVO presence. Boxplot of GFAP according to the ASPECTS (B), stratified by LVO presence (C).

## DISCUSSION

Brain-specific blood biomarker analysis is feasible in acute stroke and shows promise as an adjunct tool in acute stroke. In our study, serum GFAP levels are markedly higher in acute ICH than AIS using modern ultrasensitive immunoassays, consistent with previous published studies.^(7)^ Furthermore, we observed trends toward higher GFAP concentrations with more extensive ischemic injury on neuroimaging as well as time of stroke onset. These findings, although preliminary, lend credence to the potential role of brain-specific biomarkers as adjunct tools to evaluate patient eligibility to acute stroke treatments. This concept is of particular interest given emerging portable point-of-care technology that measures GFAP levels within minutes.^(8)^

Due its small sample size, the findings are considered hypothesis-generating and require validation in larger prospective cohorts. Nevertheless, they are intriguing regarding the potential role of GFAP measurement in acute stroke including ischemic stroke of unknown onset. If confirmed in larger studies, rapid GFAP levels have the potential to serve as an important clinical adjunct tool in the out-of-hospital setting to optimise detection, diagnosis, and triage of stroke patients to appropriate levels of care.^(8)^ In particular, the recently published INTERACT 4 study provides evidence that prehospital management of undifferentiated stroke requires development of portable tools to improve diagnosis and management.^(9)^

## CONCLUSION

In acute stroke, serum GFAP shows potential as an adjunct tool for the distinction between ICH and AIS. Specific to AIS, GFAP may also offer insight into time from onset, and extent of ischemic tissue injury on neuroimaging, particularly in LVO stroke. These preliminary findings merit further study.

## Data Availability

Data is available by the corresponding author upon reasonable request

## ACKNOWLEDGMENTS

JP: First draft of the manuscript. LCG: Study design, study procedures oversight. LCG, CD, PC: Sample manipulation, data acquisition. AD, CL: SIMOA analyses. All authors: Critical review of the manuscript.

